# Recognition and Treatment of Primary Aldosteronism in the Updated Guideline Era

**DOI:** 10.64898/2026.06.08.26355219

**Authors:** Cheng-Hsuan Tsai, Yu-Ching Chang, Chin-Chen Chang, Wan-Chen Wu, Yi-Yao Chang, Uei-Lin Chen, Bo-Ching Lee, Chi-Sheng Hung, Kuo-How Huang, Jeff Shih-Chieh Chueh, Vin-Cent Wu, Yen-Hung Lin

**Affiliations:** Division of Cardiology, Department of Internal Medicine, National Taiwan University Hospital and National Taiwan University College of Medicine, Taipei, Taiwan; Primary Aldosteronism Center at National Taiwan University Hospital, Taipei, Taiwan; Cardiovascular Center and Division of Cardiology, Department of Internal Medicine, National Taiwan University Hospital Hsin-Chu Branch, Hsin-Chu, Taiwan; Department and Graduate Institute of Forensic Medicine, National Taiwan University College of Medicine, Taipei, Taiwan; Department of Medical Imaging, National Taiwan University Hospital and National Taiwan University College of Medicine, Taipei, Taiwan; Division of Endocrinology and Metabolism, Department of Internal Medicine, National Taiwan University Hospital and National Taiwan University College of Medicine, Taipei, Taiwan; Cardiology Division of Cardiovascular Medical Center, Far Eastern Memorial Hospital, New Taipei City, Taiwan; Graduate Institute of Medicine, Yuan Ze University, Taoyuan, Taiwan; Department of Internal Medicine, Min-Sheng General Hospital, Taoyuan, Taiwan; Department of Urology, National Taiwan University Hospital and National Taiwan University College of Medicine, Taipei, Taiwan; Division of Nephrology, Department of Internal Medicine, National Taiwan University Hospital and National Taiwan University College of Medicine, Taipei, Taiwan

**Keywords:** hypertension, primary aldosteronism, aldosterone, renin, diagnsis

## Abstract

**Background:** Primary aldosteronism (PA) is increasingly recognized as a common cause of hypertension. The 2025 Endocrine Society guideline introduced a simplified diagnostic framework, but its real-world clinical implications remain unclear.

**Methods:** We conducted a multicenter retrospective cohort study of hypertensive patients undergoing PA testing in Taiwan. PA was defined biochemically according to the 2025 Endocrine Society criteria. Multivariable logistic regression identified factors associated with PA diagnosis and aldosterone-targeted therapy. Among patients with suppressed renin (≤1 ng/mL/h), restricted cubic splines evaluated the adjusted association between renin and PA probability.

**Results:** Among 18,766 patients undergoing PA testing, 6,760 (36.0%) met diagnostic criteria for PA. PA was associated with older age, female sex, lower potassium, resistant hypertension, and a higher antihypertensive medication burden. Among patients with suppressed renin, lower renin remained significantly associated with higher adjusted PA probability. However, only 39.0% of patients with PA received aldosterone-targeted therapy, including 28.2% who received mineralocorticoid receptor antagonist therapy within 6 months and 9.4% who underwent adrenalectomy during follow-up. Lower renin, higher aldosterone, lower potassium, and resistant hypertension were associated with aldosterone-targeted therapy, while younger patients with fewer comorbidities were more likely to undergo adrenalectomy.

**Conclusions:** Using the updated diagnostic framework, PA was highly prevalent among hypertensive patients undergoing PA testing. Nevertheless, many patients who met these biochemical criteria did not receive aldosterone-targeted therapy in routine care. These findings highlight the potential treatment implications of broader PA recognition and support the development of practical pathways to guide MRA therapy, adrenalectomy referral, and individualized management.

## Introduction

Primary aldosteronism (PA) is one of the most common and clinically important causes of secondary hypertension and has traditionally been estimated to account for approximately 10% to 20% of patients with hypertension^1–6^. Compared with essential hypertension, PA is associated with a higher burden of cardiovascular, renal, and metabolic complications because renin-independent aldosterone secretion promotes sodium retention, volume expansion, vascular injury, inflammation, and target-organ damage beyond the effect of blood pressure alone^7–10^. Despite its clinical importance, PA remains substantially underrecognized in routine practice^11–15^, in part because diagnostic pathways have traditionally required multistep testing, confirmatory testing, and specialist interpretation.

The 2025 Endocrine Society guideline introduced an updated and simplified diagnostic framework for PA, emphasizing the combination of suppressed renin, elevated aldosterone, and an elevated aldosterone-to-renin ratio (ARR), without a mandatory requirement for medication withdrawal or diagnostic confirmation with aldosterone suppression testing, traditionally referred to as confirmatory testing^16^. This updated approach reflects the growing recognition that PA may represent a broader spectrum of renin-independent aldosterone excess rather than a rare binary diagnosis limited to patients with florid presentations such as hypokalemia, resistant hypertension, or adrenal tumors. However, the real-world implications of applying this updated framework to a large clinical population remain uncertain^3,17,18^. In particular, it remains unclear how frequently patients undergoing PA testing would meet the updated biochemical criteria and which clinical features are associated with PA under this updated diagnostic framework.

Beyond diagnosis, an equally important question is how frequently patients with biochemical evidence of PA receive disease-specific treatment. Mineralocorticoid receptor antagonist (MRA) therapy and adrenalectomy are established aldosterone-targeted treatment strategies, but their use in routine practice may be limited by clinical inertia, diagnostic uncertainty, comorbidities, surgical candidacy, and variation in referral pathways^19,20^. The 2025 Endocrine Society guideline suggests more liberal use of MRA therapy in patients with PA. However, the gap between the expected high use of aldosterone-targeted therapy under the 2025 Endocrine Society guideline and previous clinical practice remains unclear. Applying the updated framework to real-world data provides an opportunity to estimate the burden of biochemically defined PA and to evaluate the gap between biochemical recognition and aldosterone-targeted treatment.

Therefore, using this retrospective multicenter hospital-based cohort in Taiwan, we applied the updated 2025 Endocrine Society criteria to hypertensive patients who underwent PA testing. We aimed to determine the prevalence and clinical features associated with PA diagnosis based on the 2025 Endocrine Society guideline, and to quantify the frequency of aldosterone-targeted therapy, including MRA therapy and adrenalectomy, among patients meeting the diagnostic criteria for PA. This study aimed to clarify both the potential clinical burden of PA in the updated guideline era and the magnitude of the remaining treatment gap.

## Methods

### Study population

We identified all adult patients with hypertension who underwent PA testing, defined as measurement of both plasma renin activity and plasma aldosterone concentration, in the National Taiwan University Hospital–integrated Medical Database between January 1, 2006, and December 31, 2022. The database is maintained and curated by National Taiwan University Hospital using standardized data management procedures to ensure data accuracy and completeness^21,22^. For patients with multiple renin measurements, the first available plasma renin activity measurement during the study period was used as the index test. Hypertension was defined by diagnostic codes for hypertension and/or documented use of antihypertensive medications. Patients younger than 18 years of age and those without an available plasma aldosterone concentration measurement were excluded. The final study population included 18,766 patients who underwent PA testing with paired renin and aldosterone measurements. This study was approved by the Research Ethics Committee of National Taiwan University Hospital.

### Definition of Primary Aldosteronism

PA was defined biochemically according to the 2025 Endocrine Society clinical practice guideline. Patients were classified as having PA if they had suppressed renin, defined as plasma renin activity ≤1.0 ng/mL/h, together with plasma aldosterone concentration ≥ 10 ng/dL and an ARR >20 ng/dL per ng/mL/h^16^. Patients who underwent PA testing but did not meet these biochemical criteria were classified as non-PA. Plasma renin activity and plasma aldosterone concentration were measured using immunoassays as part of routine clinical care across participating hospitals.

### Covariates

Baseline characteristics were collected at the time of the index renin test, including age, sex, body mass index, laboratory measurements, comorbidities, cardiovascular history, and antihypertensive medication use. Laboratory measurements included plasma renin activity, plasma aldosterone concentration, ARR, serum creatinine, estimated glomerular filtration rate, sodium, potassium, fasting glucose, and alanine aminotransferase. Comorbidities and cardiovascular history were identified using International Classification of Diseases, Ninth and Tenth Revision codes (**Supplemental Table 1**) and included diabetes mellitus, hyperlipidemia, coronary artery disease, atrial fibrillation, obstructive sleep apnea, chronic kidney disease, heart failure, acute myocardial infarction, intracranial hemorrhage, and ischemic stroke. Baseline antihypertensive medication use included renin-angiotensin system inhibitors, beta-blockers, alpha-blockers, vasodilators, thiazide diuretics, calcium channel blockers, MRAs, and other antihypertensive agents, including clonidine and minoxidil. Resistant hypertension was operationally defined as the use of at least 4 antihypertensive medication classes. Hypokalemia was defined as serum potassium <3.5 mEq/L. Young-onset hypertension was operationally defined as age <40 years at the index test, because the exact age at hypertension diagnosis was not available in the database.

### Aldosterone-Targeted Therapy

Aldosterone-targeted therapy was defined as receipt of MRA therapy or adrenalectomy after the index test. Because spironolactone is the only MRA reimbursed by Taiwan’s National Health Insurance program, MRA therapy was defined as spironolactone use in this study. We separately evaluated MRA prescription within 6 months after the index test and any MRA prescription recorded during follow-up. Adrenalectomy was identified using procedure codes during follow-up. Any aldosterone-targeted therapy was defined as receipt of either MRA therapy or adrenalectomy from the index test through the end of follow-up.

### Statistical Analysis

Baseline characteristics were summarized according to PA status. Continuous variables were presented as mean ± standard deviation or median with interquartile range, as appropriate, and categorical variables were presented as frequency and percentage. Multivariable logistic regression models were used to identify factors associated with biochemical PA diagnosis. The fully adjusted model included age, sex, body mass index, serum creatinine, serum potassium, diabetes mellitus, chronic kidney disease, coronary artery disease, resistant hypertension, hypokalemia, young-onset hypertension, obstructive sleep apnea, history of ischemic stroke, history of heart failure, and baseline use of renin-angiotensin system inhibitors, beta-blockers, diuretics, and mineralocorticoid receptor antagonists. As a sensitivity analysis, we evaluated the prevalence of PA across major baseline medication-use groups, including renin-angiotensin system inhibitor users, beta-blocker users, thiazide users, and patients without interfering medications. No interfering medications was defined as no baseline use of renin-angiotensin system inhibitors, beta-blockers, or thiazide diuretics.

Among patients meeting biochemical criteria for PA, fully adjusted multivariable logistic regression models were used to evaluate factors associated with MRA prescription within 6 months after the index test and adrenalectomy during follow-up. These models included the same clinical covariates, with additional adjustment for log-transformed plasma renin activity and log-transformed plasma aldosterone concentration. Among patients with suppressed renin, defined as plasma renin activity ≤1.0 ng/mL/h, restricted cubic spline models were used to evaluate the adjusted association between plasma renin activity and the probability of meeting biochemical criteria for PA. Plasma renin activity was log-transformed before spline modeling. Restricted cubic spline analyses were performed using R software, version 4.5.3, with the rms package, and other statistical analyses were performed using IBM SPSS Statistics, version 25.0. A two-sided P value <0.05 was considered statistically significant.

## Results

### Study Population and Prevalence of PA

A total of 19,448 patients with hypertension underwent PA testing between January 1, 2006, and December 31, 2022. After excluding 91 patients younger than 18 years of age and 591 patients without available aldosterone measurements, 18,766 patients were included in the final analysis. Using the 2025 Endocrine Society criteria for PA, 6,760 patients (36.0%) were classified as having PA and 12,006 patients (64.0%) were classified as non-PA. The baseline characteristics of the study population are shown in **Table 1**. Patients with PA were older than those without PA (57.8 ± 14.7 vs 54.4 ± 17.0 years) and were less likely to be male (45.2% vs 54.6%). Baseline comorbidities were generally comparable between the PA and non-PA groups, although diabetes mellitus, chronic kidney disease, and heart failure were slightly less frequent among patients with PA. As expected, patients with PA had lower plasma renin activity, higher plasma aldosterone concentration, higher ARR, and lower serum potassium levels than patients without PA. Median plasma renin activity was 0.3 ng/mL/h in the PA group and 1.9 ng/mL/h in the non-PA group, while median aldosterone-to-renin ratio was 90.2 and 11.5, respectively.

**Table 1.** Demographic and clinical characteristics of patients receiving plasma renin activity examination.

| Variable | Total<br>18766 | PA<br>(*n* = 6760) | Non-PA<br>(*n* =12006) |
| --- | --- | --- | --- |
| Age, year | 55.6 ± 16.3 | 57.8 ± 14.7 | 54.4 ±17.0 |
| Age >65 years | 5629 (30.0) | 2197 (32.5) | 3432 (28.6) |
| Male sex | 9607 (51.2) | 3054 (45.2) | 6553 (54.6) |
| Body weight, kg | 68.8 ± 15.7 | 67.5 ± 14.9 | 69.5 ± 16.0 |
| Body height, cm | 163.0 ± 9.0 | 162.1±8.7 | 163.5±9.1 |
| Body mass index, kg/m<sup>2</sup> | 25.7 ± 4.6 | 25.5±4.4 | 25.8±4.7 |
| Biochemical studies |  |  |  |
| Plasma renin activity | 1.2 (0.4, 2.7) | 0.3 (0.1, 0.6) | 1.9 (1.3, 4.9) |
| Plasma aldosterone concentration | 25.2 (15.3, 37.5) | 26.8 (18.2, 37.6) | 24.0 (13.5, 37.5) |
| Aldosterone-to-renin ratio | 20.0 (9.0, 58.4) | 90.2 (45.6, 241.6) | 11.5 (6.1, 18.9) |
| Hemoglobin, g/dL | 13.2 ± 2.3 | 13.2±2.4 | 13.2±2.4 |
| Serum creatinine, mg/dL | 1.1 ± 0.9 | 1.1±0.8 | 1.2±0.9 |
| eGFR, mL/min/1.73m<sup>2</sup> | 82.5 ± 31.5 | 82.7±29.8 | 82.4±32.5 |
| Sodium, mEq/L | 138.8 ±3.7 | 139.3±3.6 | 138.5±3.8 |
| Potassium, mEq/L | 4.1 ±0.6 | 4.0±0.6 | 4.1±0.6 |
| Fasting glucose, mg/dL | 107.3 ±30.2 | 105.7±27.8 | 108.3±31.5 |
| ALT, U/L | 26.6 ± 22.0 | 25.6±20.6 | 27.2±22.7 |
| Comorbidity |  |  |  |
| Diabetes mellitus | 3257 (17.4) | 1081 (16.0) | 2176 (18.1) |
| Hyperlipidemia | 3809 (20.4) | 1341 (19.8) | 2468 (20.6) |
| Coronary artery disease | 1738 (9.3) | 634 (9.4) | 1104 (9.2) |
| Atrial fibrillation | 429 (2.3) | 158 (2.3) | 271 (2.3) |
| Obstructive sleep apnea | 569 (3.0) | 218 (3.2) | 351 (2.9) |
| Chronic kidney disease | 2140 (11.4) | 698 (10.3) | 1442 (12.0) |
| Heart failure | 544 (2.9) | 146 (2.2) | 398 (3.3) |
| Acute myocardial infarction | 173 (0.9) | 44 (0.7) | 129 (1.1) |
| Intracranial hemorrhage | 541 (2.9) | 164 (2.4) | 377 (3.1) |
| Ischemic stroke | 1106 (5.9) | 381 (5.6) | 725 (6.0) |
| Anti-hypertensive medications at baseline |  |  |  |
| RASi | 9873 (52.6) | 3430 (50.7) | 6443 (53.7) |
| Beta-blocker | 6815 (36.3) | 2583 (38.2) | 4232 (35.2) |
| Alpha-blocker | 3829 (20.4) | 1597 (23.6) | 2232 (18.6) |
| Vasodilator | 1178 (6.3) | 391 (5.8) | 787 (6.6) |
| Thiazide | 2118 (11.3) | 734 (10.9) | 1384 (11.5) |
| Calcium channel blocker | 10932 (58.3) | 4081 (60.4) | 6851 (57.1) |
| Others (Clonidine, Minoxidil) | 603 (3.2) | 242 (3.6) | 361 (3.0) |
| MRA | 870 (4.6) | 329 (4.9) | 541 (4.5) |
| Number of medications | 2.0±1.5 | 2.0±1.5 | 1.95±1.5 |

**Abbreviation:** ALT, alanine aminotransferase; eGFR, estimated glomerular filtration rate; HbA1c, glycated hemoglobin; MRA, mineralocorticoid receptor antagonist; RASi, renin-angiotensin system inhibitors.
Data were presented as frequency (percentage), mean ± standard deviation or median [25<sup>th</sup>, 75<sup>th</sup> percentiles].

In sensitivity analyses stratified by baseline medication use, the prevalence of biochemical PA was similar across major medication-use groups, including renin-angiotensin system inhibitor users, beta-blocker users, thiazide users, and patients without interfering medications (**Figure 2**).

**Figure 1.**
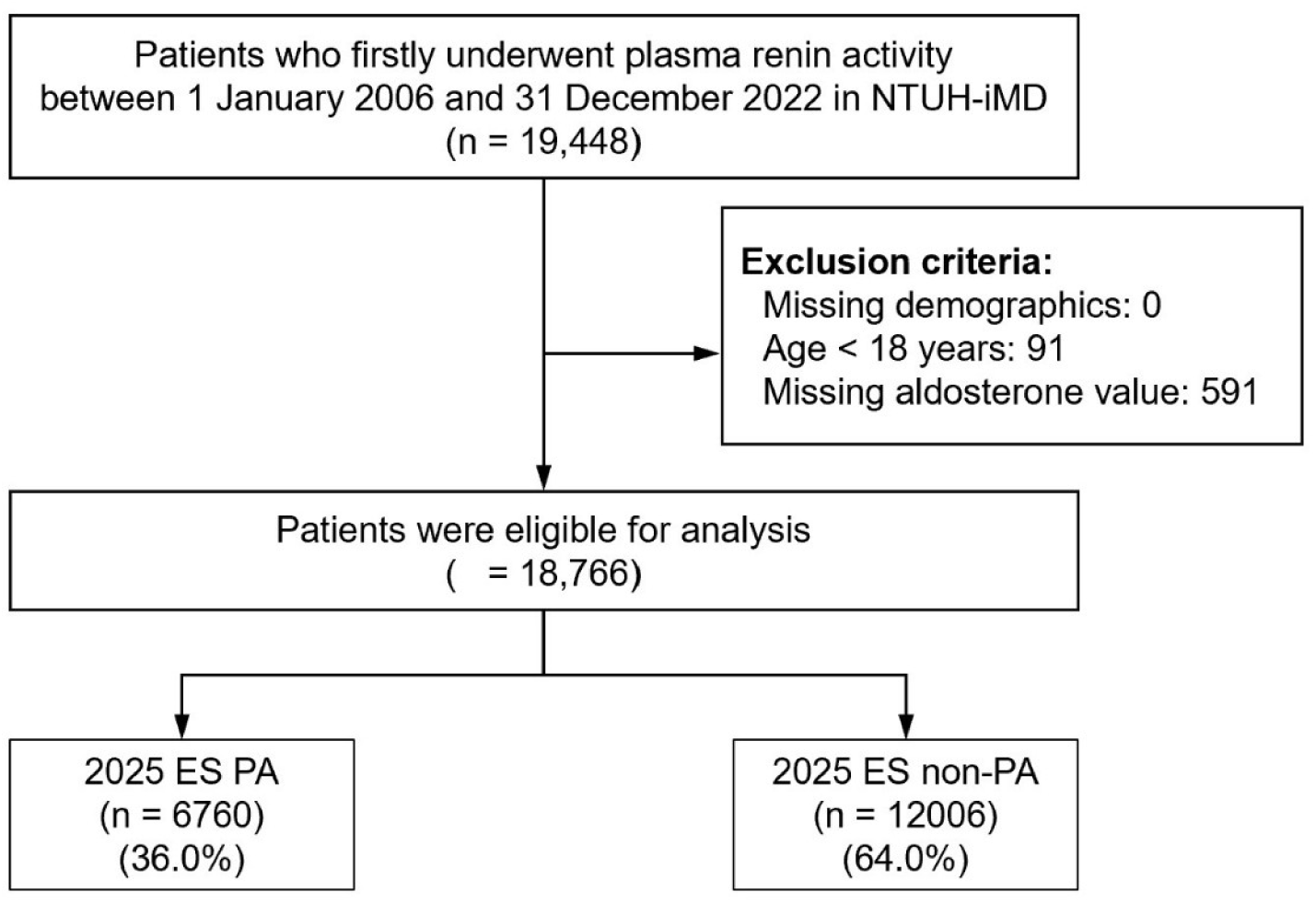
Study flow chart Flow chart of patient selection from the National Taiwan University Hospital-integrated Medical Database. ES, endocrine society; NTUH-iMD, National Taiwan University Hospital-integrated Medical Database

**Figure 2.**
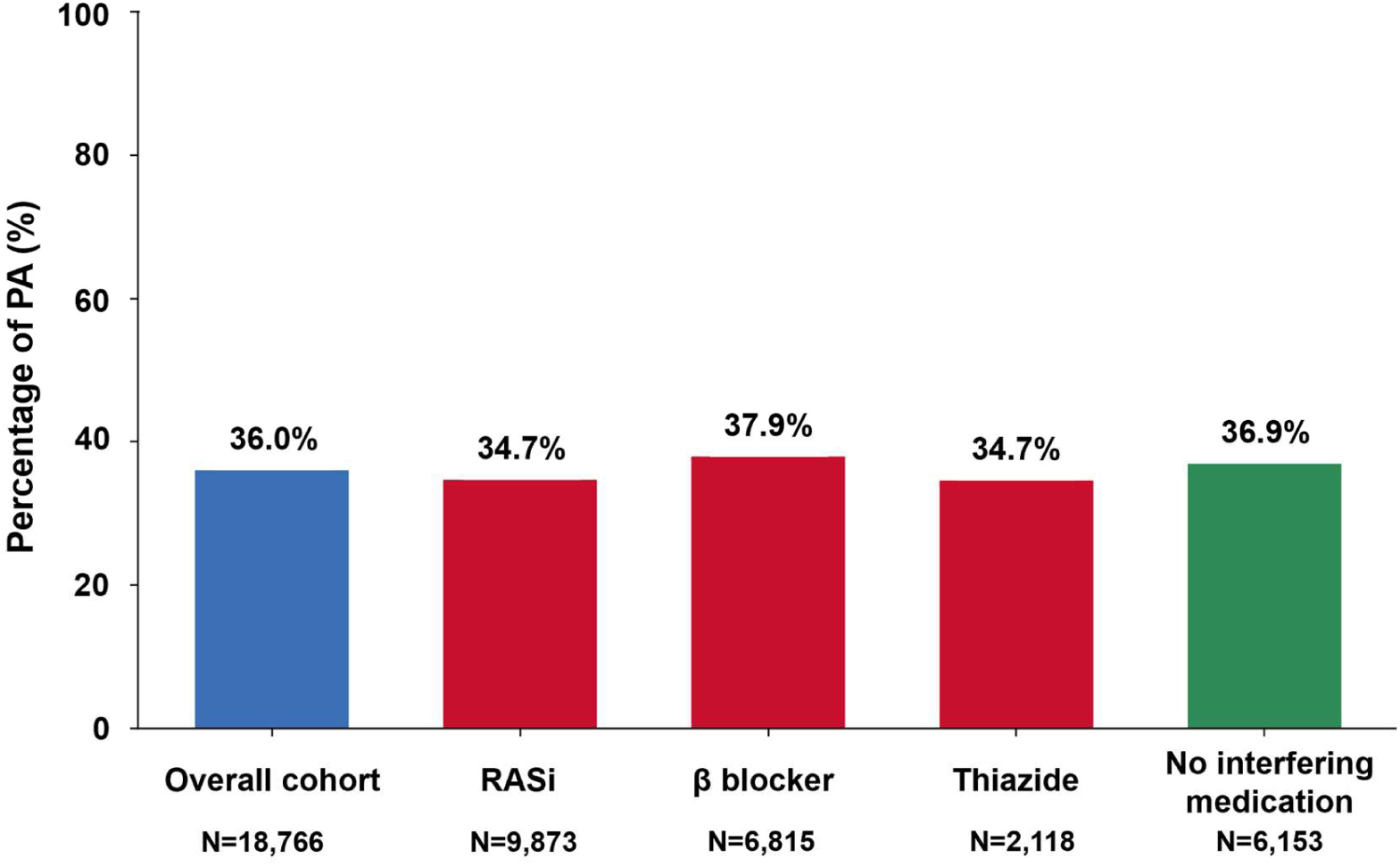
PA prevalence by baseline medication-use group. Prevalence of PA among the overall cohort and according to baseline medication-use groups. The prevalence of PA was 36.0% in the overall cohort, 34.7% among renin-angiotensin system inhibitor users, 37.9% among beta-blocker users, 34.7% among thiazide users, and 36.9% among patients without interfering medications. No interfering medications was defined as no baseline use of renin-angiotensin system inhibitors, beta-blockers, or thiazide diuretics. PA, primary aldosteronism. RASi, renin-angiotensin system inhibitors

### Clinical Factors Associated with the diagnosis of PA

The clinical factors associated with PA are listed in **Table 2**. In fully adjusted logistic regression models, older age, female sex, lower serum potassium, resistant hypertension, and baseline beta-blocker use were independently associated with higher odds of PA. Resistant hypertension was associated with 22% higher odds of PA (OR, 1.224; 95% CI, 1.092–1.373), whereas higher serum potassium was associated with lower odds of PA (OR, 0.826; 95% CI, 0.762–0.896). In contrast, diabetes mellitus, history of heart failure, young-onset hypertension, baseline renin–angiotensin system inhibitor use, and diuretic use were associated with lower odds of PA. Baseline MRA use was not associated with PA diagnosis after multivariable adjustment. Among patients with suppressed renin, the adjusted probability of PA increased progressively as plasma renin activity decreased (**Figure 3**). Restricted cubic spline analysis demonstrated both significant linear and non-linear associations between plasma renin activity and PA probability among patients with plasma renin activity ≤1.0 ng/mL/h (both P <0.001).

**Figure 3.**
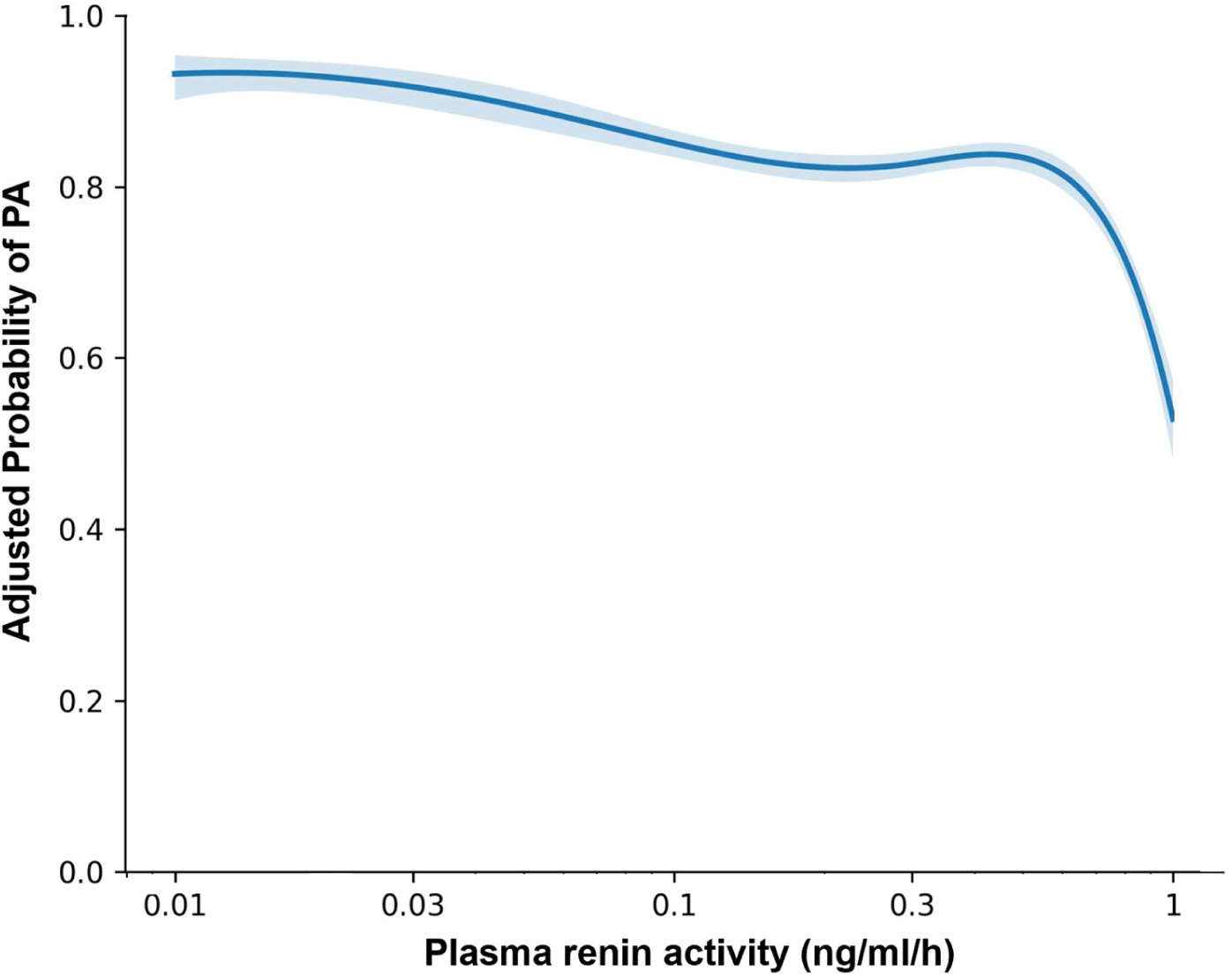
Adjusted probability of PA across suppressed renin levels. Adjusted probability of PA across plasma renin activity levels among patients with suppressed renin, defined as plasma renin activity ≤1.0 ng/mL/h. Plasma renin activity was modeled using restricted cubic splines after log transformation. The model was adjusted for age, sex, renal function, body mass index, baseline antihypertensive medication use, and antihypertensive medication burden. The x-axis is displayed on a logarithmic scale with values shown in original units. PA, primary aldosteronism.

**Table 2.** Clinical factors associated with the diagnosis of primary aldosteronism.

|  | Univariable logistic regression analysis |  | Multivariable logistic regression analysis |  |
| --- | --- | --- | --- | --- |
|  | OR (95% CI) | P value | OR (95% CI) | P value |
| Age, year | 1.013 (1.011 – 1.015) | <0.001 | 1.006 (1.003 – 1.009) | <0.001 |
| Sex, Male | 0.686 (0.646 – 0.728) | <0.001 | 0.758 (0.707 – 0.812) | <0.001 |
| Body mass index, kg/m<sup>2</sup> | 0.987 (0.980 – 0.994) | <0.001 | 1.004 (0.996 – 1.012) | 0.289 |
| Serum creatinine, mg/dL | 0.892 (0.860 – 0.925) | <0.001 | 0.947 (0.905 – 0.990) | 0.017 |
| Potassium, mEq/L | 0.796 (0.753 – 0.842) | <0.001 | 0.826 (0.762 – 0.896) | <0.001 |
| Diabetes Mellitus | 0.860 (0.794 – 0.931) | <0.001 | 0.785 (0.714 – 0.862) | <0.001 |
| Chronic kidney disease | 0.844 (0.767 – 0.928) | <0.001 | 0.896 (0.794 – 1.011) | 0.075 |
| Coronary artery disease | 1.022 (0.922 – 1.132) | 0.678 | 0.980 (0.873 – 1.100) | 0.728 |
| Resistant hypertension | 1.087 (1.003 – 1.178) | 0.043 | 1.224 (1.092 – 1.373) | 0.001 |
| Hypokalemia | 1.381 (1.261 – 1.512) | <0.001 | 1.050 (0.920 – 1.198) | 0.471 |
| Young-onset hypertension | 0.483 (0.444 – 0.526) | <0.001 | 0.570 (0.502 – 0.648) | <0.001 |
| Obstructive sleep apnea | 1.106 (0.932 – 1.314) | 0.248 | 1.046 (0.869 – 1.260) | 0.634 |
| History of heart failure | 0.929 (0.818 – 1.056) | 0.261 | 0.884 (0.771 – 1.014) | 0.078 |
| History of ischemic stroke | 0.644 (0.531 – 0.780) | <0.001 | 0.594 (0.484 – 0.730) | <0.001 |
| RASi | 0.889 (0.838 – 0.944) | <0.001 | 0.857 (0.798 – 0.922) | <0.001 |
| Beta blocker | 1.136 (1.068 – 1.208) | <0.001 | 1.142 (1.056 – 1.235) | 0.001 |
| Diuretics | 0.935 (0.850 – 1.028) | 0.164 | 0.819 (0.727 – 0.922) | 0.001 |
| Previous MRA use | 1.084 (0.942 – 1.248) | 0.259 | 1.000 (0.857 – 1.166) | 0.997 |

**Abbreviation:** MRA, mineralocorticoid receptor antagonist.

**Table 3.** Clinical and biochemical factors associated with aldosterone-targeted therapy among patients with PA.

|  | Multivariable logistic regression analysis |  |  |  |
| --- | --- | --- | --- | --- |
|  | MRA use within 6 months after testing |  | Receiving adrenalectomy |  |
|  | OR (95% CI) | P value | OR (95% CI) | P value |
| Age, year | 1.003 (0.997 – 1.009) | 0.335 | 0.967 (0.958 – 0.976) | <0.001 |
| Sex, Male | 0.893 (0.777 – 1.025) | 0.109 | 1.124 (0.920 – 1.374) | 0.254 |
| Body mass index, kg/m<sup>2</sup> | 1.022 (1.006 – 1.038) | 0.006 | 1.007 (0.985 – 1.029) | 0.559 |
| Serum creatinine, mg/dL | 0.686 (0.614 – 0.768) | <0.001 | 0.702 (0.561 – 0.877) | 0.002 |
| Potassium, mEq/L | 0.558 (0.471 – 0.662) | <0.001 | 0.630 (0.487 – 0.816) | <0.001 |
| Log plasma renin activity | 0.694 (0.612 – 0.786) | <0.001 | 0.678 (0.574 – 0.799) | <0.001 |
| Log plasma aldosterone concentration | 22.301 (16.662 – 28.849) | <0.001 | 9.983 (6.871 – 14.506) | <0.001 |
| Diabetes Mellitus | 1.005 (0.831 – 1.214) | 0.962 | 0.831 (0.606 – 1.138) | 0.249 |
| Chronic kidney disease | 1.075 (0.836 – 1.381) | 0.575 | 0.582 (0.358 – 0.944) | 0.028 |
| Coronary artery disease | 0.768 (0.610 – 0.967) | 0.024 | 0.878 (0.600 – 1.284) | 0.501 |
| Resistant hypertension | 2.169 (1.744 – 2.697) | <0.001 | 1.453 (1.049 – 2.012) | 0.025 |
| Hypokalemia | 1.388 (1.081 – 1.781) | 0.010 | 1.962 (1.394 – 2.763) | <0.001 |
| Young-onset hypertension | 0.885 (0.683 – 1.148) | 0.358 | 1.291 (1.024 – 1.629) | <0.001 |
| Obstructive sleep apnea | 1.044 (0.728 – 1.497) | 0.814 | 1.089 (0.607 – 1.951) | 0.776 |
| History of heart failure | 1.870 (1.209 – 2.894) | 0.005 | 0.721 (0.255 – 2.043) | 0.538 |
| History of ischemic stroke | 0.537 (0.399 – 0.723) | <0.001 | 0.172 (0.075 – 0.391) | <0.001 |
| Renin-Angiotensin-System inhibitor | 1.059 (0.919 – 1.221) | 0.425 | 0.788 (0.644 – 0.964) | 0.020 |
| Beta blocker | 1.240 (1.066 – 1.442) | 0.005 | 0.815 (0.654 – 1.015) | 0.068 |
|  | MRA use within 6 months after testing |  | Receiving adrenalectomy |  |
|  | OR (95% CI) | P value | OR (95% CI) | P value |
| Diuretics | 0.940 (0.748 – 1.183) | 0.599 | 0.754 (0.524 – 1.084) | 0.128 |
| Previous MRA use | 14.716 (10.183 – 21.267) | <0.001 | 1.624 (1.145 – 2.302) | 0.007 |

**Abbreviation:** MRA, mineralocorticoid receptor antagonist.

### Aldosterone-Targeted Therapy Among Patients With PA

Among patients meeting biochemical criteria for PA, aldosterone-targeted therapy was used in fewer than half of patients during follow-up. MRA therapy was prescribed within 6 months after testing in 28.2% of patients, and 36.7% received an MRA prescription at any time during follow-up. Adrenalectomy was performed in 9.4% of patients, and 39.0% received either MRA therapy or adrenalectomy during follow-up (**Figure 4**).

**Figure 4.**
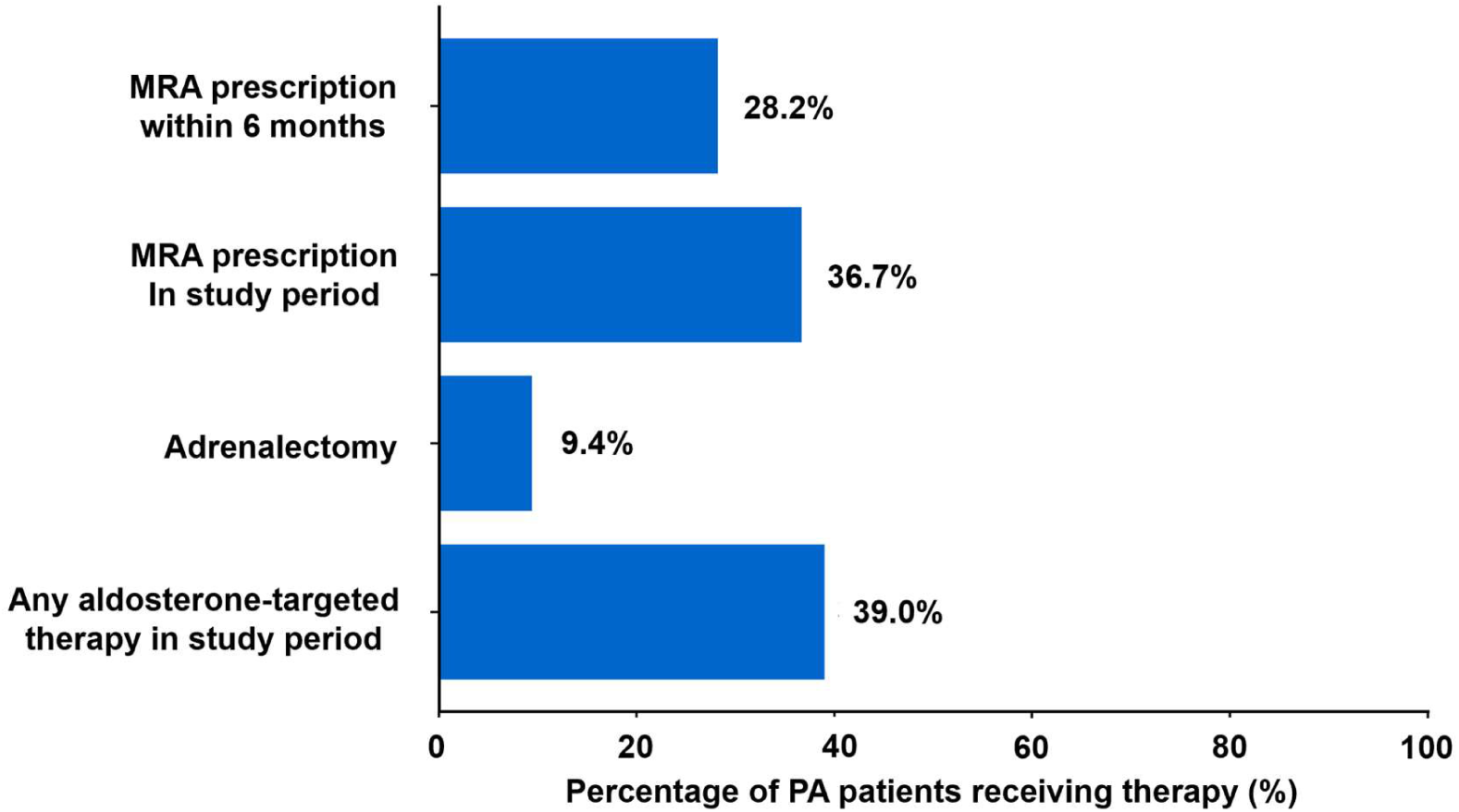
Aldosterone-targeted therapy among patients with PA. Proportions of patients meeting biochemical criteria for PA who received aldosterone-targeted therapy after the index test. MRA therapy within 6 months, any MRA prescription during follow-up, adrenalectomy, and any aldosterone-targeted therapy during follow-up are shown. Any aldosterone-targeted therapy was defined as receipt of either MRA therapy or adrenalectomy from the index test through the end of follow-up. MRA, mineralocorticoid receptor antagonist; PA, primary aldosteronism.

In multivariable logistic regression models, lower plasma renin activity, higher plasma aldosterone concentration, lower serum potassium, higher body mass index, resistant hypertension, history of heart failure, baseline beta-blocker use, and previous MRA use were associated with higher odds of MRA prescription within 6 months after testing. Resistant hypertension was strongly associated with MRA use within 6 months (OR, 2.169; 95% CI, 1.744–2.697). Higher plasma aldosterone concentration was also strongly associated with MRA use (OR, 22.301; 95% CI, 16.662–28.849), whereas higher serum potassium and higher plasma renin activity were associated with lower odds of MRA use.

Factors associated with adrenalectomy were generally similar to those associated with MRA use, although adrenalectomy was more strongly associated with younger age and lower comorbidity burden. Younger age, lower serum creatinine, lower serum potassium, lower plasma renin activity, higher plasma aldosterone concentration, resistant hypertension, hypokalemia, young-onset hypertension, and previous MRA use were associated with higher odds of receiving adrenalectomy during follow-up. In contrast, chronic kidney disease, history of ischemic stroke, and baseline renin–angiotensin system inhibitor use were associated with lower odds of adrenalectomy, suggesting that age and comorbidity burden may have influenced surgical treatment selection.

## Discussion

In this multicenter hospital-based cohort of hypertensive patients undergoing PA testing, application of the updated 2025 Endocrine Society diagnostic algorithm identified PA in more than one-third of tested patients with hypertension. Older age, female sex, lower potassium, and resistant hypertension were associated with higher odds of PA, whereas several baseline antihypertensive medications were associated with PA classification in directions consistent with their known effects on renin and aldosterone measurements^16^. However, the prevalence of PA remained high and was similar across baseline antihypertensive medication-use groups. Among patients with suppressed renin, lower plasma renin activity was progressively associated with a higher adjusted probability of PA. Despite meeting biochemical criteria for PA, aldosterone-targeted therapy was not universally used, with only 39.0% of patients receiving MRA therapy or adrenalectomy during follow-up. These findings suggest that the updated diagnostic framework may identify a substantial group of patients with biochemical evidence of renin-independent aldosterone excess who could potentially benefit from aldosterone-targeted therapy, highlighting the need for clinicians to recognize this population and assess their suitability for such targeted treatment.

Historically, the diagnosis of PA has required a multistep approach, beginning with ARR-based screening, followed by medication adjustment or withdrawal, confirmatory aldosterone suppression testing, adrenal imaging, and in selected patients, adrenal venous sampling^23–25^. These requirements can create practical barriers to PA testing and may contribute to low screening rates in routine practice^11,13,15,26^. Although this approach might improve diagnostic specificity, its complexity may contribute to underdiagnosis and delayed treatment in routine practice^18,27–29^. The updated 2025 Endocrine Society guideline represents a more pragmatic diagnostic framework by emphasizing the biochemical pattern of suppressed renin, elevated aldosterone, and elevated ARR, without requiring medication withdrawal or routine confirmatory aldosterone suppression testing in all patients^16^. In this context, our findings illustrate the clinical impact of applying this updated framework to real-world PA testing: more than one-third of tested hypertensive patients met biochemical criteria for PA. Importantly, this suggests that broader biochemical recognition may identify a substantial group of patients who would previously have been classified or managed as non-PA hypertension, despite biochemical evidence of renin-independent aldosterone excess.

Another important finding of this study is the graded relationship between renin suppression and the probability of PA. Among patients with suppressed renin, lower plasma renin activity was progressively associated with a higher adjusted probability of meeting PA criteria. This finding supports the concept that renin suppression should not be interpreted only as a binary screening threshold, but also as a biological marker reflecting the degree of renin-independent aldosterone excess. Patients with very low renin may represent a group with more pronounced sodium retention, volume expansion, and mineralocorticoid receptor activation, whereas those with mildly suppressed renin may have a less severe PA pathophysiology. The clinical relevance of renin may also extend beyond diagnosis. Prior studies have shown that suppressed renin is associated with higher cardiovascular risk, and that patients with PA or low-renin may experience better outcomes when MRA therapy increases renin, suggesting more complete mineralocorticoid receptor blockade^30^. In this context, renin may serve not only as a diagnostic tool for identifying renin-independent aldosterone excess, but also as a potential marker of disease severity, treatment adequacy, and long-term risk^31–33^. Our findings are consistent with this broader role of renin and support the emerging view that PA exists along a continuum of disease spectrum rather than as a strictly dichotomous disease state^17,18,34^.

We also demonstrated the relatively limited use of aldosterone-targeted therapy among patients meeting 2025 Endocrine Society criteria for PA. In our cohort, 28.2% of patients received MRA therapy within 6 months after testing, 36.7% received MRA therapy during follow-up, 9.4% underwent adrenalectomy, and 39.0% received either MRA therapy or adrenalectomy. The current guideline suggests a more liberal use of MRA in patients with PA^35^. Our results demonstrated a substantial gap between our previous clinical practice and current guideline suggestions. MRA therapy and adrenalectomy are established treatment strategies for PA^36,37^, but treatment selection depends on multiple factors, including biochemical severity, blood pressure burden, renal function, comorbidities, surgical candidacy, patient preference, referral patterns, and whether patients are clinically recognized as having PA.

In our analysis, lower renin, higher aldosterone, lower potassium, and resistant hypertension were associated with greater likelihood of aldosterone-targeted therapy, suggesting that clinicians were more likely to treat patients with more overt biochemical or clinical features. However, patients with less florid presentations may still have clinically relevant renin-independent aldosterone excess under the updated framework. Therefore, broader biochemical recognition of PA should be accompanied by practical strategies to determine which patients are suitable for MRA therapy, adrenalectomy evaluation, or longitudinal biochemical monitoring.

Several limitations should be acknowledged. First, this was a retrospective observational study, and residual confounding and selection bias cannot be excluded. Because our cohort included patients who underwent PA testing within a multicenter hospital-based health system, the observed prevalence of PA should not be generalized to all patients with hypertension. However, the database included medical centers and affiliated regional hospitals across different geographic regions in Taiwan, which may partially mitigate single-center referral bias and improve the relevance of our findings to real-world PA testing practice. Second, PA was defined retrospectively according to the updated 2025 Endocrine Society criteria using renin and aldosterone measurements obtained in routine clinical practice. Medication withdrawal and sampling conditions were not standardized, which may have led to false-positive or false-negative biochemical classification in some patients. However, subgroup analyses according to baseline medication use showed only minimal differences in PA proportions across medication strata, suggesting that medication-related misclassification was unlikely to fully explain the main findings. Third, resistant hypertension was based on the number of antihypertensive medication classes rather than blood pressure control status, because standardized blood pressure measurements were not available. Similarly, young-onset hypertension was operationally defined using age at the index test rather than the exact age at hypertension diagnosis. These operational definitions may have led to misclassification and may not fully identify all patients who met guideline-defined resistant hypertension or true young-onset hypertension.

### Perspectives

In this study, application of the updated 2025 Endocrine Society framework to a large real-world PA testing cohort identified PA in more than one-third of tested hypertensive patients. Lower renin within the suppressed range was associated with progressively higher PA probability, supporting a biological gradient of renin-independent aldosterone excess. However, aldosterone-targeted therapy was not universally used among patients meeting biochemical criteria. These findings highlight the need for clinicians to recognize patients with biochemical evidence of PA and to systematically assess their suitability for MRA therapy, adrenalectomy evaluation, or longitudinal biochemical monitoring.

## Disclosure of interest

All other coauthors have nothing to disclose.

## Funding

CHT was supported by Taiwan Society of Cardiology, the National Science and Technology Council, Taiwan grant 113-2314-B-002-152-MY2 and National Taiwan University Hospital grant 113-M0023, 114-M0030.

## Data Availability

The data underlying this article will be shared on reasonable request to the corresponding author.

## Acknowledgements

We thank the staff of the Eighth and Second Core Lab in the Department of Medical Research at National Taiwan University Hospital for technical support. We also thank all members of the TAIPAI Study Group for help during the study. We would like to express our thanks to the staff of Department of Medical Research for providing clinical data from National Taiwan University Hospital-integrative Medical Data Center. The authors also thank Alfred Hsing-Fen Lin, Zoe Ya-Zhu Syu, and Winnie Su-Hui Wu, who served in Raising Statistics Consultant Inc., for their assistance with statistical analysis.

## Nonstandard Abbreviation and Acronyms

ARR: aldosterone-to-renin ratio
MRA: mineralocorticoid receptor antagonist
PA: primary aldosteronism
PAC: plasma aldosterone concentration

## Novelty and Relevance

### What Is New

- Applying the updated 2025 Endocrine Society diagnostic framework identified PA in more than one-third of hypertensive patients undergoing primary aldosteronism testing. Despite meeting biochemical criteria for PA, fewer than half of patients with PA received aldosterone-targeted therapy during follow-up.
- Among patients with suppressed renin, lower plasma renin activity was progressively associated with higher PA probability, supporting a biological gradient of renin-independent aldosterone excess.

### What Is Relevant

- PA is more broadly recognized under the updated diagnostic framework than under traditional multistep diagnostic pathways.
- Despite meeting biochemical criteria, fewer than half of patients received aldosterone-targeted therapy, suggesting a gap between biochemical recognition and disease-specific treatment.

### Clinical Implications

- Clinicians should pay closer attention to patients with suppressed renin and biochemical evidence of renin-independent aldosterone excess. Practical pathways are needed to assess suitability for mineralocorticoid receptor antagonist therapy, adrenalectomy evaluation, or longitudinal biochemical monitoring in patients with PA.

---

**Supplemental Table 1.**
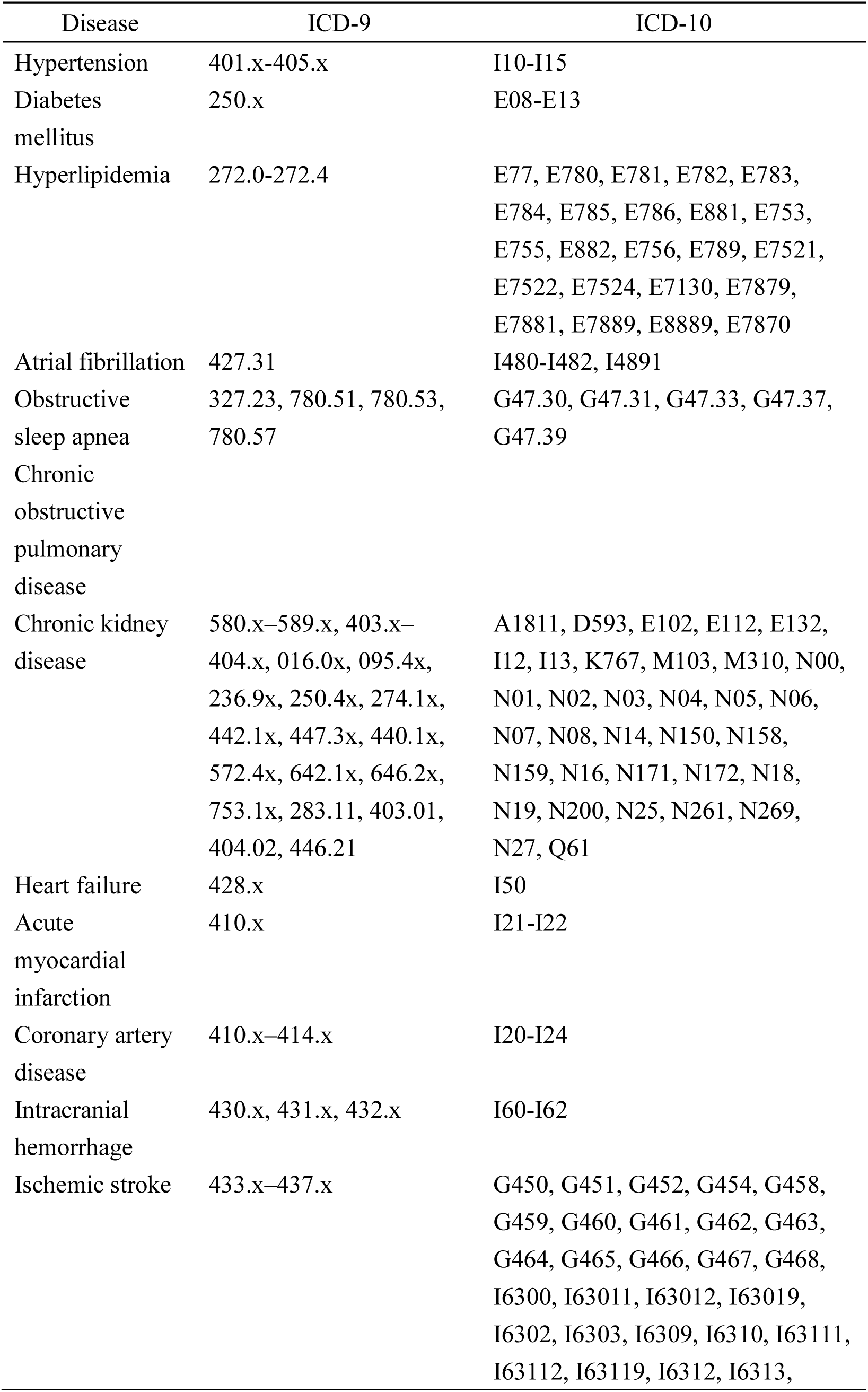
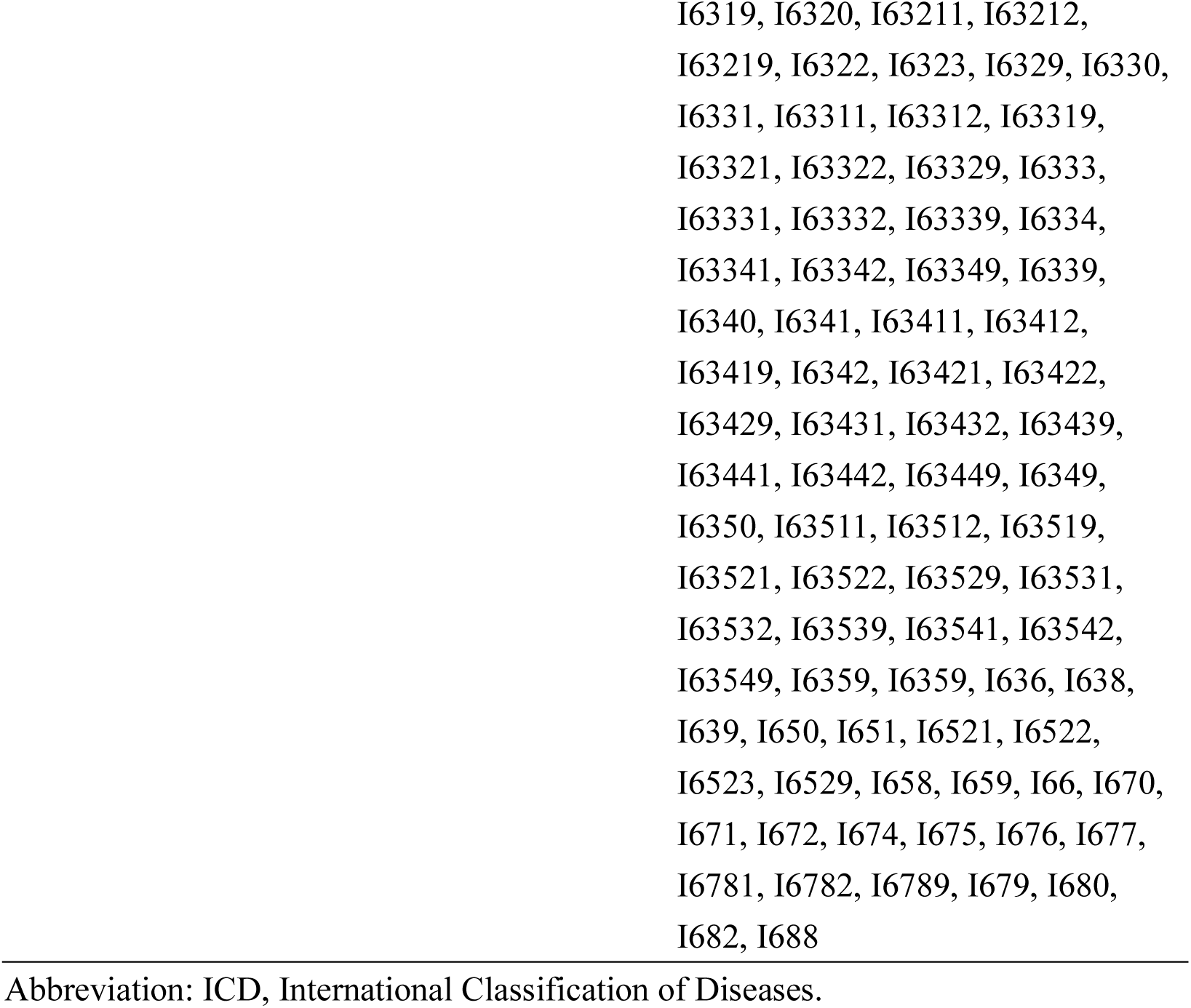
ICD diagnostic codes used in the study.

| Disease | ICD-9 | ICD-10 |
| --- | --- | --- |
| Hypertension | 401.x-405.x | I10-I15 |
| Diabetes mellitus | 250.x | E08-E13 |
| Hyperlipidemia | 272.0-272.4 | E77, E780, E781, E782, E783, E784, E785, E786, E881, E753, E755, E882, E756, E789, E7521, E7522, E7524, E7130, E7879, E7881, E7889, E8889, E7870 |
| Atrial fibrillation | 427.31 | I480-I482, I4891 |
| Obstructive sleep apnea | 327.23, 780.51, 780.53, 780.57 | G47.30, G47.31, G47.33, G47.37, G47.39 |
| Chronic obstructive pulmonary disease |  |  |
| Chronic kidney disease | 580.x–589.x, 403.x–404.x, 016.0x, 095.4x, 236.9x, 250.4x, 274.1x, 442.1x, 447.3x, 440.1x, 572.4x, 642.1x, 646.2x, 753.1x, 283.11, 403.01, 404.02, 446.21 | A1811, D593, E102, E112, E132, I12, I13, K767, M103, M310, N00, N01, N02, N03, N04, N05, N06, N07, N08, N14, N150, N158, N159, N16, N171, N172, N18, N19, N200, N25, N261, N269, N27, Q61 |
| Heart failure | 428.x | I50 |
| Acute myocardial infarction | 410.x | I21-I22 |
| Coronary artery disease | 410.x–414.x | I20-I24 |
| Intracranial hemorrhage | 430.x, 431.x, 432.x | I60-I62 |
| Ischemic stroke | 433.x–437.x | G450, G451, G452, G454, G458, G459, G460, G461, G462, G463, G464, G465, G466, G467, G468, I6300, I63011, I63012, I63019, I6302, I6303, I6309, I6310, I63111, I63112, I63119, I6312, I6313, |

I6319, I6320, I63211, I63212, I63219, I6322, I6323, I6329, I6330, I6331, I63311, I63312, I63319, I63321, I63322, I63329, I6333, I63331, I63332, I63339, I6334, I63341, I63342, I63349, I6339, I6340, I6341, I63411, I63412, I63419, I6342, I63421, I63422, I63429, I63431, I63432, I63439, I63441, I63442, I63449, I6349, I6350, I63511, I63512, I63519, I63521, I63522, I63529, I63531, I63532, I63539, I63541, I63542, I63549, I6359, I6359, I636, I638, I639, I650, I651, I6521, I6522, I6523, I6529, I658, I659, I66, I670, I671, I672, I674, I675, I676, I677, I6781, I6782, I6789, I679, I680, I682, I688
Abbreviation: ICD, International Classification of Diseases.

## Notes

### Competing Interest Statement

The authors have declared no competing interest.

### Author Declarations

This retrospective cohort study was approved by the Research Ethics Committee of National Taiwan University Hospital. The requirement for written informed consent was waived because the study used retrospectively collected clinical data from the National Taiwan University Hospital-integrated Medical Database and involved no direct patient contact.

